# Association of Free Triiodothyronine with Long-Term Prognosis in Hypertrophic cardiomyopathy patients with HFpEF: A Dual center research study

**DOI:** 10.1101/2023.04.18.23288777

**Authors:** Xiangbin Meng, Jing-Jia Wang, Jun Gao, Kuo Zhang, Jun Wen, Xu-Liang Wang, Yuan-Geng-Shuo Wang, Ji-Lin Zheng, Yu-Peng Liu, Jing-Jing Song, Jie Yang, Yi-Tian Zheng, Chen Li, Wen-Yao Wang, Chunli Shao, Yi-Da Tang

## Abstract

**Objective:** The objective of this study was to identify prognostic factors in Hypertrophic cardiomyopathy patients with HFpEF, with a particular focus on the role of FT3 levels.

**Research Design and Methods:** We conducted a retrospective cohort study of 992 patients with HFpEF who were admitted to our two medical centers between 2009 and 2019. We collected demographic and clinical data, including FT3 levels, and conducted univariate and multivariate Cox analyses, KM survival curve analysis, and RSC curve analysis to identify prognostic factors and evaluate the non-linear predictive value of FT3.

**Results:** We found that age, atrial fibrillation, and NT-proBNP levels were all significant prognostic factors in patients with heart failure. Additionally, FT3 levels were a significant independent predictor of all-cause mortality and cardiac transplantation. Patients with lower FT3 levels had worse long-term prognoses, and the critical value of FT3 was 2.885.

**Conclusions:** Our findings suggest that FT3 levels are an important prognostic factor in patients with heart failure and should be considered when evaluating patient outcomes. Clinicians should monitor FT3 levels and consider interventions to maintain or improve thyroid function in patients with heart failure.

**Article Highlights:**

- FT3 levels are a significant independent predictor of all-cause mortality and cardiac transplantation in Hypertrophic cardiomyopathy patients with HFpEF
- Patients with hypertrophic cardiomyopathy combined with HFpEF have a poorer long-term prognosis with lower FT3 levels, with a critical FT3 value of 2.885
- Age, atrial fibrillation, and NT-proBNP levels were also significant prognostic factors in Hypertrophic cardiomyopathy patients with HFpEF
- Clinicians should monitor FT3 levels and consider interventions to maintain or improve thyroid function in Hypertrophic cardiomyopathy patients with HFpEF.

## 1. Introduction

Heart failure (HF) is a chronic and progressive condition in which the heart is unable to pump enough blood to meet the body’s needs. It is a major cause of morbidity and mortality worldwide, with an estimated prevalence of 1-2% in the general population. The prognosis of HF is highly variable, and identifying prognostic factors is critical for guiding clinical decision-making and improving patient outcomes. The importance of identifying prognostic factors in HF cannot be overstated. Such factors can help clinicians stratify patients according to their risk of adverse outcomes, tailor treatment plans to individual patients, and monitor disease progression over time. However, despite extensive research in this area, there is still a need to identify new prognostic markers that can improve risk prediction and guide treatment decisions.

HF with preserved ejection fraction (HFpEF) is a specific type of HF in which the ejection fraction, or the percentage of blood that is pumped out of the heart with each beat, is normal or only slightly reduced. Patients with HFpEF typically present with symptoms such as shortness of breath, fatigue, and exercise intolerance, and the prevalence of HFpEF is increasing due to the aging population and the high prevalence of comorbidities such as hypertension and diabetes.

Hypertrophic cardiomyopathy (HCM) is a genetic disorder characterized by abnormal thickening of the heart muscle, which can lead to various clinical manifestations including arrhythmias, heart failure, and sudden cardiac death. Previous studies have shown that the prevalence of HFpEF is high in patients with HCM, and HFpEF has been shown to be an independent predictor of adverse outcomes in this population.

Thyroid hormones are known to have important effects on the cardiovascular system, including the regulation of heart rate, contractility, and vascular tone. Thyroid hormone profile acts as a fundamental regulator in cardiovascular homeostasis in both physiological conditions and pathological processes ^[1,2]^. Abnormal thyroid hormone levels, such as hypothyroidism or hyperthyroidism, have been associated with various cardiovascular diseases including HF. Our previous study, together with other clinical evidence, indicated that thyroid hormone levels, particularly the level of free triiodothyronine (FT3), correlate with exacerbated cardiac function ^[3-6]^ and hypothyroid status was an independent risk factor of adverse events in patients with cardiovascular diseases. The potential mechanism underlying the influence of thyroid hormone levels on cardiovascular disease involves the regulation of gene expression and signaling pathways involved in cardiac hypertrophy, fibrosis, and inflammation^[7,8]^. FT3 is a thyroid hormone that plays a critical role in regulating cardiac function, and previous studies have suggested that low FT3 levels are associated with poor outcomes in patients with HF^[8]^. However, the relationship between FT3 and prognosis of patients with HCM and HFpEF is still not well understood, and further research is needed to elucidate this relationship and identify optimal cutoff values for risk stratification.

Given the high prevalence of HFpEF in patients with HCM and the potential role of thyroid hormones in cardiovascular disease, there is a need to identify diagnostic factors that may help predict outcomes in this population. In this study, we aimed to investigate the prognostic value of thyroid hormone levels in patients with HCM and HFpEF, and to explore the potential mechanism underlying this association.

## 2. Research Design and Methods

### 2.1 Ethics statement

The study was in accordance with the ethical guidelines of the declaration of Helsinki and China’s regulations and guidelines on good clinical practice and was approved by the Ethics Committee of the Peking University Third Hospital. Before the start of the study, we received written informed consents from all participants.

### 2.2 Design, patients, and outcome measure

#### 2.2.1 Study Patients

This study was a two-center, retrospective cohort study. All patients in this study were evaluated at two medical centers: the Peking University Third Hospital, and Fuwai Hospital (National Center of Cardiovascular Diseases, China). Between October 1, 2009, and December 31, 2019, a total of 2802 patients (age≥16 years) were diagnosed with HOCM. Among them, 187 patients with EF<50% were excluded from this study. In the end, 992 patients with hypertrophic cardiomyopathy diagnosed with HFpEF were included in this study. Patients were divided into three groups according to the FT3 levels: Tertile 1: (FT3<2.77 pg/mL, n=336), Tertile 2 :(2.77 pg/mL≤ FT3 ≤ 3.09 pg/mL, n=335), Tertile 3 (FT3>3.10 pg/mL, n=321).

#### 2.2.2 Diagnosis of HOCM

The diagnosis of HOCM was based on ^[9-11]^: 1, wall thickness ≥15 mm in one or more LV myocardial segments, as measured by any imaging technique (echocardiography, cardiac magnetic resonance imaging, or computed tomography); 2, wall thickness (13-14mm) with family history, non-cardiac symptoms and signs, electrocardiogram(ECG) abnormalities, laboratory tests, and muti-modality cardiac imaging; 3, patients with LV outflow tract obstruction (LVOTO) were based on: dynamic LVOT obstruction due to systolic anterior motion of the mitral valve, with an LVOT gradient ≥30 mmHg at rest or during physiological provocation such as Valsalva maneuver, standing, and exercise. Significant dynamic LVOT obstruction was documented by 2-dimensional and Doppler echocardiography or, in those cases where echocardiography was insufficient, by invasive hemodynamic catheterization with provocation.

#### 2.2.3 Diagnosis of HFpEF

In our study, the diagnosis of HFpEF was determined by evaluating a combination of symptoms, signs, left ventricular ejection fraction (LVEF), and concentrations of NT-proBNP. Specifically, we followed the 2017 American Heart Association (AHA) criteria for diagnosis^[12]^, which categorizes patients with LVEF ≥50%, NT-proBNP ≥800 pg/mL, and symptoms or signs of heart failure as having HFpEF. Those who did not meet these criteria were classified as non-HF.

#### 2.2.4 Thyroid function test

Thyroid function was assessed after heart failure symptoms were controlled with regular oral medication prior to discharge, rather than during the acute phase. The thyroid function test was conducted at a median of 10 days after hospital admission. Blood samples were drawn after a 12-hour fast and the serum levels of thyroid hormone and thyroid stimulating hormone were measured using radioimmunoassay (Immulite 2000; Siemens, Germany).

### 2.3 Follow-up and endpoints

Follow-up started at the time of the patient’s first clinic contact after October 1, 2009. At baseline, all patients were evaluated for the following characteristics: age, sex, NYHA functional class, maximum left ventricular wall thickness (LVWT), maximum (Provo cable) LVOT gradient, left ventricular function, atrial fibrillation, and conventional risk factors for cardiac death.

The primary endpoints of this study were all-cause mortality and cardiac transplantation during long-term follow-up. Mortality and adverse events were retrieved from hospital patient records at the center where follow-up occurred, from civil service population registers, and information provided by patients themselves and/or their general practitioners. Patients lost to follow-up were censored upon the last contact with them. If no events occurred during follow-up, the administrative censoring date was set for December 31, 2019.

### 2.4 Data Analysis

Statistical analysis was assessed with SPSS 27.0 statistical package for Windows, and R software version 4.2.2 (R Foundation for Statistical Computing). All continuous variables are presented as means ± SD and analysis of variance was used to compare means across multiple groups. The relationships between parametric variables were assessed by multiple linear regression analysis. Initial differences in baseline characteristics between achieved treatment groups were sought in bivariable analysis by using χ^2^ tests, Fisher exact tests, Student *t* tests, and Kruskal-Wallis analysis of variance. Cox proportional hazards model was used to estimate the hazard ratio (HR). Kaplan-Meier analysis was used to study the cumulative survival of different groups. A P-value less than 0.05 was considered statistically significant.

Restricted cubic spline (RCS) is a statistical method used to model nonlinear relationships between a continuous predictor variable and an outcome variable. It is an extension of the linear regression model and allows for flexible modeling of the relationship by using cubic polynomial functions. In our study, we use the RCS method provides a flexible and powerful way to model nonlinear relationships in R. We chose to use four knots, placed at the 10th, 50th, and 90th percentiles of the distribution of FT3 in our study population, to allow for a flexible yet parsimonious modeling of the non-linear relationship between FT3 and cardiovascular events. We assessed the overall fit of the model using the likelihood ratio test and examined the shape of the spline function using graphical displays.

### 2.5 Data and Resource Availability Statement

The datasets generated during and/or analyzed in the current study are available from the corresponding author upon reasonable request.

## 3. Results

The study population consisted of 992 HCM patients with HFpEF, with an average age of 53.60±13.83 years. The majority of patients were male (58.2%). Atrial fibrillation was present in 22.5% of patients, while 21.0% had unexplained syncope. The mean NT-proBNP level was 2278.18±1743.44 fmol/mL, and the mean FT3 level was 2.94±0.58 pg/mL.

Table 1. It shows the baseline clinical characteristics of the study population divided into the three tertiles based on the level of FT3. The total number of participants in the study was 992, with Tertile 1 having 336 participants, Tertile 2 having 335 participants, and Tertile 3 having 321 participants. The mean age of the total population was 53.60±13.83 years, with Tertile 1 being the oldest at 57.21±13.84 years and Tertile 3 being the youngest at 50.00±13.49 years (p<0.001). Male participants made up the majority of the study population at 58.2%, with Tertile 3 having the highest proportion of male participants at 76.0% (p<0.001). There were no significant differences in body mass index (BMI), systolic and diastolic blood pressures, and LV end-diastolic diameter among the three tertiles. However, participants in Tertile 3 had a significantly higher level of FT3 than those in Tertile 1 and Tertile 2 (p<0.001). Regarding comorbidities, atrial fibrillation was more prevalent in Tertile 3 than in Tertile 1 and Tertile 2 (p<0.001). On the other hand, NYHA Class III or IV was more common in Tertile 1 than in Tertile 2 and Tertile 3 (p<0.001). In terms of medications, the use of beta-blockers, ACEI/ARB, diuretics, and calcium antagonists did not differ significantly among the three tertiles.

**Table 1.**
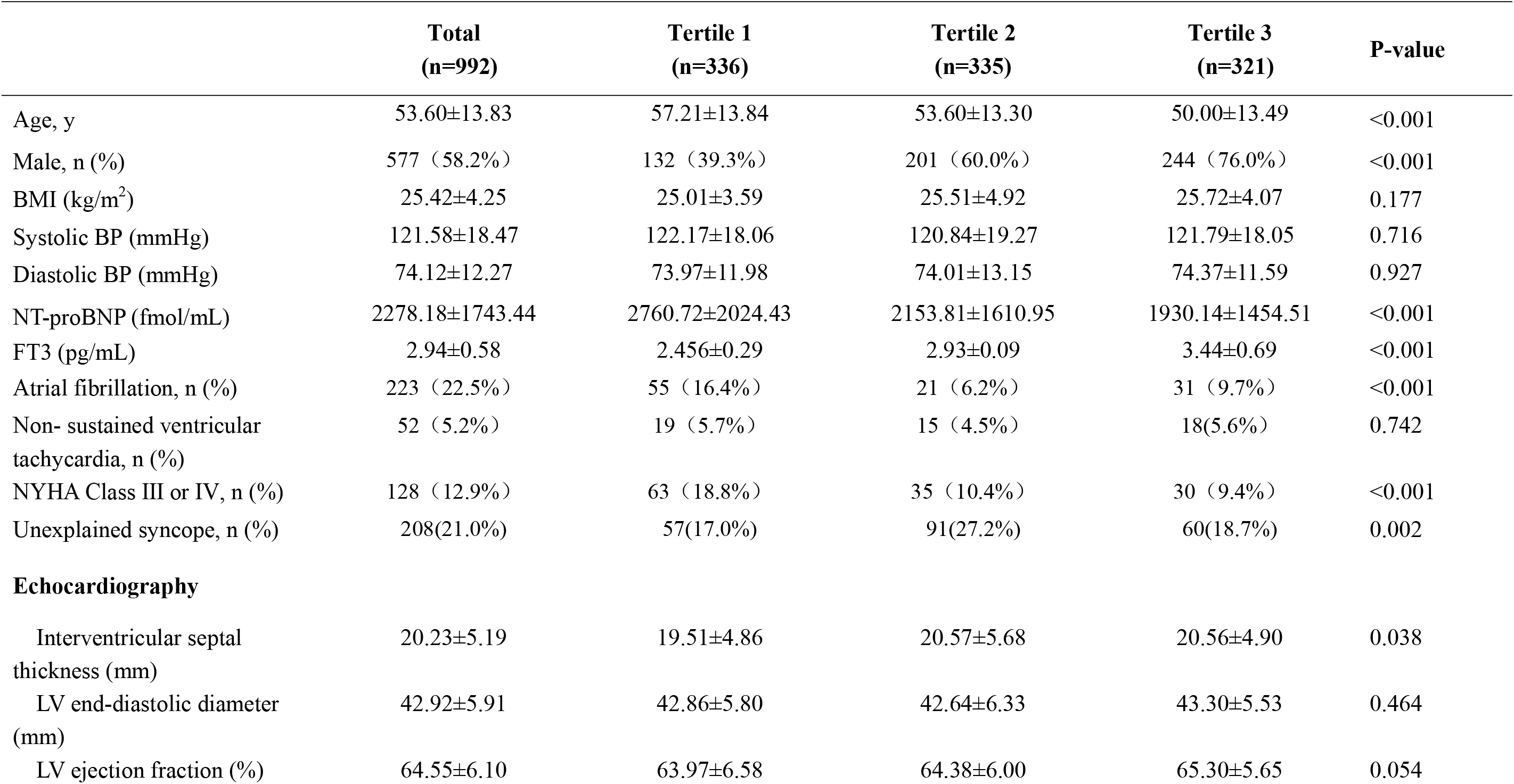
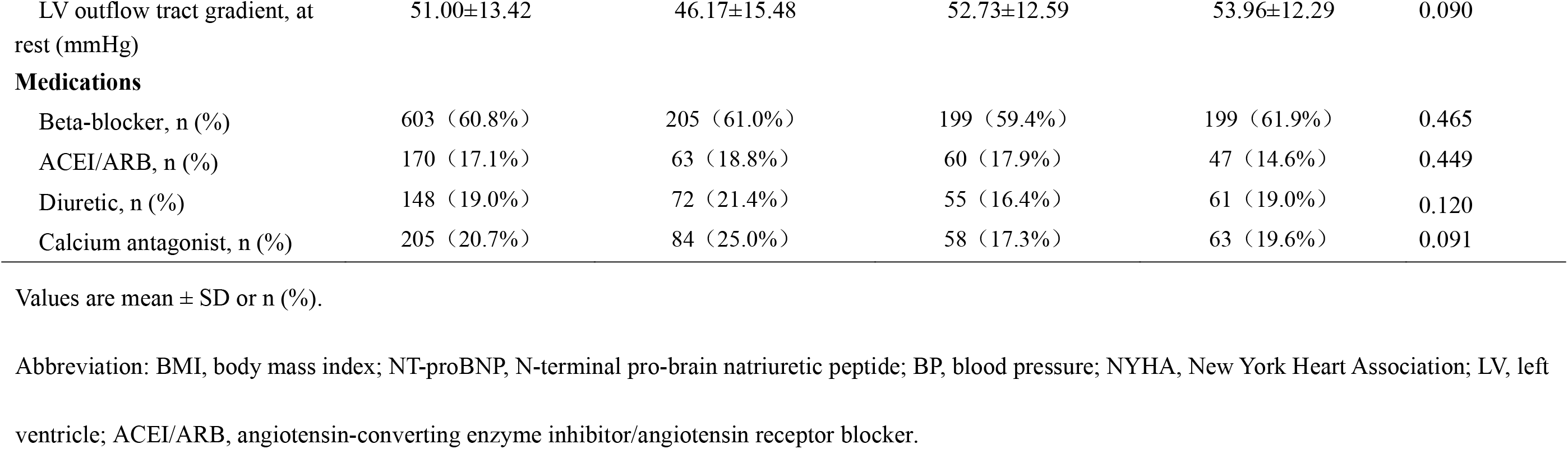
Baseline clinical characteristics of the study population divided into the three tertiles based on the level of FT3.

|  | **Total**<br>**(n=992)** | **Tertile 1**<br>**(n=336)** | **Tertile 2**<br>**(n=335)** | **Tertile 3**<br>**(n=321)** | **P-value** |
| --- | --- | --- | --- | --- | --- |
| Age, y | 53.60±13.83 | 57.21±13.84 | 53.60±13.30 | 50.00±13.49 | <0.001 |
| Male, n (%) | 577 (58.2%) | 132 (39.3%) | 201 (60.0%) | 244 (76.0%) | <0.001 |
| BMI (kg/m<sup>2</sup>) | 25.42±4.25 | 25.01±3.59 | 25.51±4.92 | 25.72±4.07 | 0.177 |
| Systolic BP (mmHg) | 121.58±18.47 | 122.17±18.06 | 120.84±19.27 | 121.79±18.05 | 0.716 |
| Diastolic BP (mmHg) | 74.12±12.27 | 73.97±11.98 | 74.01±13.15 | 74.37±11.59 | 0.927 |
| NT-proBNP (fmol/mL) | 2278.18±1743.44 | 2760.72±2024.43 | 2153.81±1610.95 | 1930.14±1454.51 | <0.001 |
| FT3 (pg/mL) | 2.94±0.58 | 2.456±0.29 | 2.93±0.09 | 3.44±0.69 | <0.001 |
| Atrial fibrillation, n (%) | 223 (22.5%) | 55 (16.4%) | 21 (6.2%) | 31 (9.7%) | <0.001 |
| Non- sustained ventricular<br>tachycardia, n (%) | 52 (5.2%) | 19 (5.7%) | 15 (4.5%) | 18(5.6%) | 0.742 |
| NYHA Class III or IV, n (%) | 128 (12.9%) | 63 (18.8%) | 35 (10.4%) | 30 (9.4%) | <0.001 |
| Unexplained syncope, n (%) | 208(21.0%) | 57(17.0%) | 91(27.2%) | 60(18.7%) | 0.002 |
| **Echocardiography** |  |  |  |  |  |
| Interventricular septal<br>thickness (mm) | 20.23±5.19 | 19.51±4.86 | 20.57±5.68 | 20.56±4.90 | 0.038 |
| LV end-diastolic diameter<br>(mm) | 42.92±5.91 | 42.86±5.80 | 42.64±6.33 | 43.30±5.53 | 0.464 |
| LV ejection fraction (%) | 64.55±6.10 | 63.97±6.58 | 64.38±6.00 | 65.30±5.65 | 0.054 |
| LV outflow tract gradient, at rest (mmHg) | 51.00±13.42 | 46.17±15.48 | 52.73±12.59 | 53.96±12.29 | 0.090 |
| **Medications** |  |  |  |  |  |
| Beta-blocker, n (%) | 603 (60.8%) | 205 (61.0%) | 199 (59.4%) | 199 (61.9%) | 0.465 |
| ACEI/ARB, n (%) | 170 (17.1%) | 63 (18.8%) | 60 (17.9%) | 47 (14.6%) | 0.449 |
| Diuretic, n (%) | 148 (19.0%) | 72 (21.4%) | 55 (16.4%) | 61 (19.0%) | 0.120 |
| Calcium antagonist, n (%) | 205 (20.7%) | 84 (25.0%) | 58 (17.3%) | 63 (19.6%) | 0.091 |

Values are mean ± SD or n (%).
Abbreviation: BMI, body mass index; NT-proBNP, N-terminal pro-brain natriuretic peptide; BP, blood pressure; NYHA, New York Heart Association; LV, left ventricle; ACEI/ARB, angiotensin-converting enzyme inhibitor/angiotensin receptor blocker.

Table 2 shows the results of the univariate Cox analysis for all-cause mortality and cardiac transplantation. The hazard ratios (HR) and 95% confidence intervals (CI) are reported for each variable, along with the corresponding p-value. The results indicate that higher levels of FT3 are associated with a lower risk of all-cause mortality and cardiac transplantation, with a HR of 0.206 and a p-value of less than 0.001. Older age is associated with a higher risk of mortality and transplantation, with a HR of 1.085 and a p-value of less than 0.001. Male gender does not appear to be significantly associated with the risk of mortality and transplantation, with a HR of 0.593 and a p-value of 0.108. Higher levels of NT-proBNP are associated with a higher risk of mortality and transplantation, with a HR of 1.026 and a p-value of less than 0.001. A history of atrial fibrillation is associated with a significantly higher risk of mortality and transplantation, with a HR of 4.240 and a p-value of less than 0.001. A history of unexplained syncope does not appear to be significantly associated with the risk of mortality and transplantation, with a HR of 1.460 and a p-value of 0.287. The interventricular septal thickness does not appear to be significantly associated with the risk of mortality and transplantation, with a HR of 1.001 and a p-value of 0.979.

**Table 2.** Univariate Cox Analysis for All-Cause Mortality and Cardiac transplantation

|  | HR (95% CI) | P value |
| --- | --- | --- |
| FT3 (pg/mL) | 0.206 (0.106, 0.402) | <0.001 |
| Age, y | 1.085(1.053, 1.118) | <0.001 |
| Male, n (%) | 0.593(0.314, 1.122) | 0.10.8 |
| NT-proBNP (per 100 fmol/mL) | 1.026(1.015, 1.037) | <0.001 |
| Atrial fibrillation, n (%) | 4.240 (2.235, 8.044) | <0.001 |
| Unexplained syncope, n (%) | 1.460(0.724, 2.944) | 0.290 |
| Interventricular septal thickness (mm) | 1.001 (0.948, 1.056) | 0.980 |

Abbreviations are listed in Table 1.

In Table 3, two different models are presented: Model 1 and Model 2. Both models are multivariate Cox regression models that examine the association between various factors and all-cause mortality and cardiac transplantation. In Model 1, the factors included age, atrial fibrillation, NT-proBNP (per 100 fmol/mL), and FT3 (pg/mL). The hazard ratio (HR) for age was 1.062 (95% CI: 1.029, 1.096), indicating that for every one year increase in age, the risk of all-cause mortality and cardiac transplantation increased by 6.2%. The HR for atrial fibrillation was 2.142 (95% CI: 1.084, 4.234), indicating that patients with atrial fibrillation had a 2.1 times higher risk of all-cause mortality and cardiac transplantation compared to those without atrial fibrillation. The HR for NT-proBNP was 1.021 (95% CI: 1.007, 1.035), indicating that for every 100 fmol/mL increase in NT-proBNP, the risk of all-cause mortality and cardiac transplantation increased by 2.1%. The HR for FT3 was 0.374 (95% CI: 0.178, 0.788), indicating that for every 1 pg/mL increase in FT3, the risk of all-cause mortality and cardiac transplantation decreased by 62.6%. In Model 2, the same factors were included as in Model 1, but FT3 was analyzed as a categorical variable (tertiles). The reference group was FT3 Tertial 3, and the HR for FT3 Tertial 2 was 0.480 (95% CI: 0.214, 1.074), indicating that patients in the second tertile of FT3 levels had a 52.0% lower risk of all-cause mortality and cardiac transplantation compared to those in the reference group. The HR for FT3 Tertial 1 was 0.336 (95% CI: 0.114, 0.988), indicating that patients in the first tertile of FT3 levels had a 66.4% lower risk of all-cause mortality and cardiac transplantation compared to those in the reference group. In addition, the HR for age, atrial fibrillation, and NT-proBNP remained significant in Model 2. The HR for age was 1.040 (95% CI: 1.002, 1.079), indicating that for every one year increase in age, the risk of all-cause mortality and cardiac transplantation increased by 4.0%. The HR for atrial fibrillation was 3.399 (95% CI: 1.356, 8.517), indicating that patients with atrial fibrillation had a 3.4 times higher risk of all-cause mortality and cardiac transplantation compared to those without atrial fibrillation. The HR for NT-proBNP was 1.023 (95% CI: 1.005, 1.041), indicating that for every 100 fmol/mL increase in NT-proBNP, the risk of all-cause mortality and cardiac transplantation increased by 2.3%.

**Table 3.** Multivariate Cox Analysis for All-Cause Mortality and Cardiac transplantation

|  | Model 1 |  | Model 2 |  |
| --- | --- | --- | --- | --- |
|  | HR (95% CI) | P value | HR (95% CI) | P value |
| Age, y | 1.062 (1.029, 1.096) | <0.001 | 1.040 (1.002, 1.079) | 0.037 |
| Atrial fibrillation | 2.142 (1.084, 4.234) | 0.028 | 3.399 (1.356, 8.517) | 0.009 |
| NT-proBNP<br>(per 100 fmol/ml) | 1.021 (1.007, 1.035) | 0.003 | 1.023 (1.005,1.041) | 0.012 |
| FT3 ((pg/mL) | 0.374 (0.178, 0.788) | 0.010 | -- | -- |
| FT3 Tertial 3 | -- | -- | 1 (reference) |  |
| FT3 Tertial 2 | -- | -- | 0.480 (0.214, 1.074) | 0.074 |
| FT3 Tertial 1 | -- | -- | 0.336(0.114, 0.988) | 0.048 |

Multivariate Cox model selected by a stepwise method with factors that were significant in the univariate analysis and established risk factors for prognosis (age, Atrial fibrillation , NT-proBNP). Model 1 was constructed based on inclusion of FT3 as continuous variable; Model 2 was constructed based on inclusion of FT3 as Categorical variable.

In Fig1, based on the KM survival curve, it seems that Group 1 had the worst long-term prognosis, and there was a statistically significant difference in prognosis among the three groups (log-rank test, p<0.001). It is important to note that the KM survival curve is a graphical representation of the survival probability over time, and it allows for the comparison of survival between different groups. In this case, it appears that patients with lower FT3 levels had a worse prognosis than those with higher FT3 levels.

**Fig. 1.**
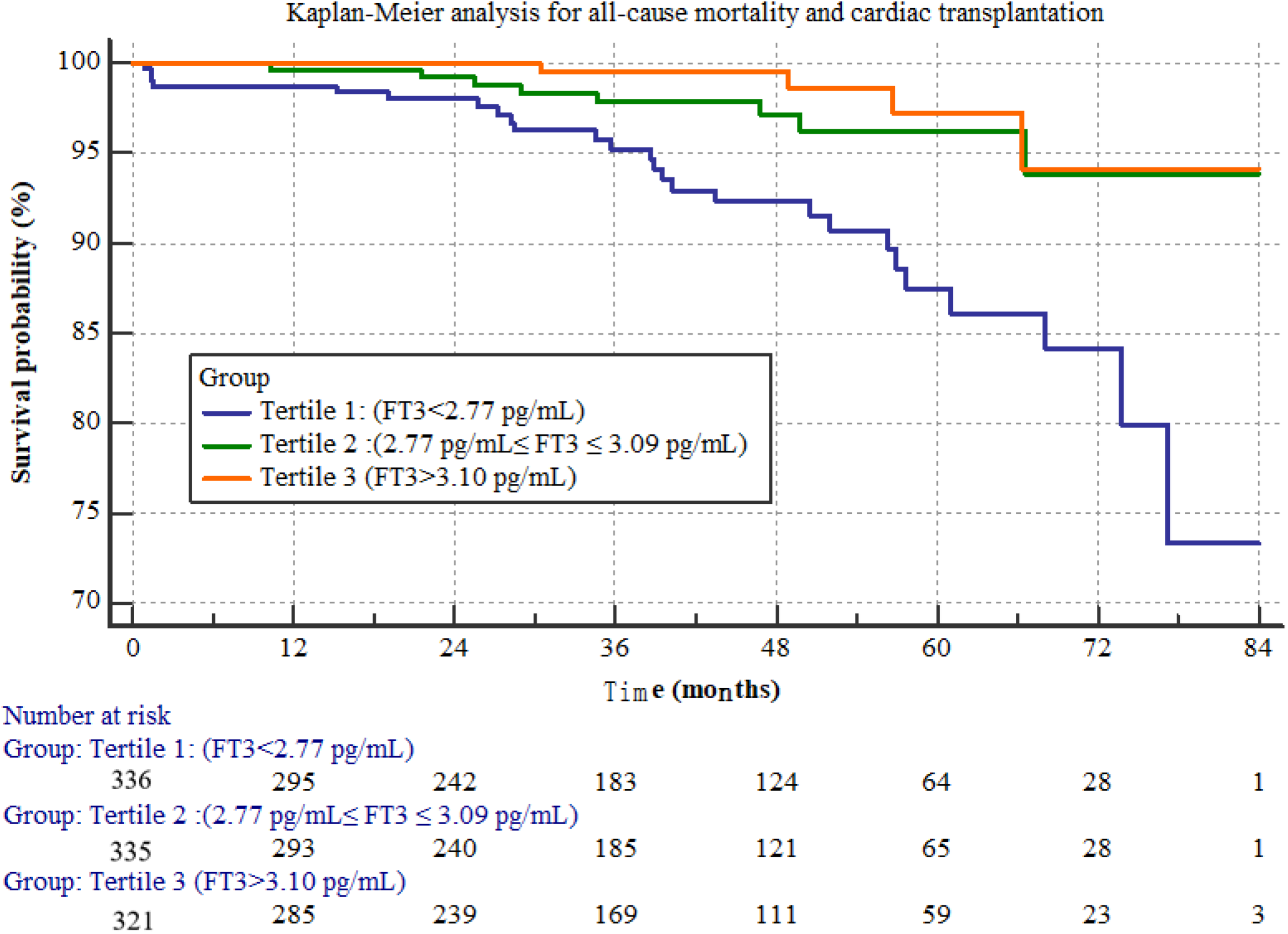

In Fig2, we used restricted cubic splines to model the relationship between the FT3 and the risk of cardiovascular events. We chose to use four knots, placed at the 10th, 50th, and 90th percentiles of the distribution of the FT3 in our study population, to allow for flexible yet parsimonious modeling of the non-linear relationship between the FT3 and cardiovascular events. We assessed the overall fit of the model using the likelihood ratio test and examined the shape of the spline function using graphical displays. The RSC curve analysis showed a non-linear relationship between FT3 levels and prognosis. The RSC curve analysis revealed an “L” shape, indicating that as the FT3 level decreased, the prognosis of the patient gradually deteriorated. The critical value of FT3 was 2.885 pg/mL.

**Fig. 2.**
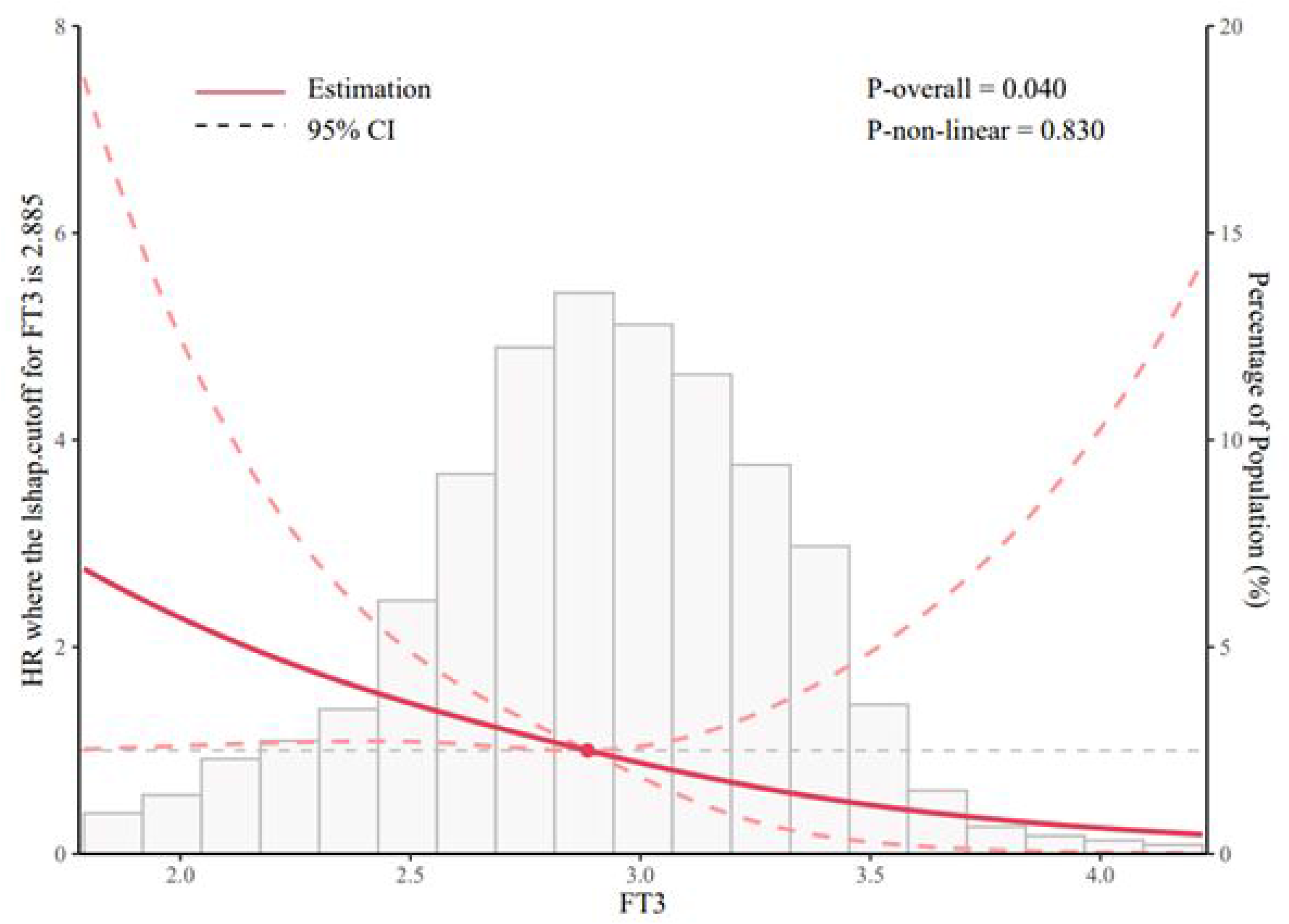
Restricted Cubic Splines (RCS) of the relationship between the FT3 and the risk of all-cause mortality and cardiac transplantation.

## 4. Discussion

Summary of the main findings This study investigated the prognostic value of FT3 in Hypertrophic cardiomyopathy patients with HFpEF. The study found that lower FT3 levels were associated with a higher risk of all-cause mortality and cardiac transplantation. The multivariate analysis showed that FT3 was an independent prognostic factor after adjusting for other clinical factors. The KM survival curve analysis showed that patients with lower FT3 levels had worse long-term prognosis. Additionally, the RSC curve analysis showed an “L” shaped relationship between FT3 levels and prognosis, with a critical value of FT3=2.885.

Comparison with previous studies Our findings are consistent with previous studies that have reported an association between low FT3 levels and poor outcomes in patients with heart failure^[13,14]^. However, our study is unique in that it used a large sample size, included various clinical factors in the analysis, and used RSC curve analysis to explore the non-linear relationship between FT3 and prognosis.

Interpretation of the results the results suggest that FT3 is a valuable prognostic factor for patients with heart failure. Lower FT3 levels are associated with worse outcomes and may be used to identify high-risk patients who require closer monitoring and more aggressive treatment. The non-linear relationship between FT3 and prognosis suggests that a critical value of FT3 may exist, below which the risk of adverse outcomes significantly increases.

Clinical implications and potential applications the identification of FT3 as a prognostic factor has important clinical implications for the management of heart failure. Clinicians should consider measuring FT3 levels in patients with heart failure to assess their risk of adverse outcomes. Additionally, our findings suggest that interventions to optimize FT3 levels may improve the prognosis of patients with heart failure.

There is growing evidence suggesting that thyroid hormones play a critical role in regulating cardiovascular function. In particular, thyroid hormone deficiency, also known as hypothyroidism, has been associated with a variety of cardiovascular diseases, including heart failure. Conversely, excess thyroid hormone, or hyperthyroidism, has also been linked to adverse cardiovascular outcomes, such as atrial fibrillation and sudden cardiac death.

The exact mechanisms by which thyroid hormones influence cardiovascular function are complex and not fully understood. It is known that thyroid hormones affect almost every aspect of cardiac function, including cardiac contractility, heart rate, and rhythm, as well as the structure and function of the blood vessels^[15,16]^. Thyroid hormones also have profound effects on the metabolism of lipids and carbohydrates, which can further impact cardiovascular health^[17]^.

One of the proposed mechanisms by which thyroid hormones affect the prognosis of heart failure is through their influence on the sympathetic nervous system^[18]^. Thyroid hormones are known to enhance sympathetic activity, which can increase heart rate and blood pressure, as well as promote cardiac hypertrophy and remodeling. These changes can ultimately lead to the development and progression of heart failure^[19]^.

Another possible mechanism is the effect of thyroid hormones on the renin-angiotensin-aldosterone system (RAAS), which is a key regulator of blood pressure and fluid balance in the body^[18]^. Thyroid hormones have been shown to stimulate the RAAS, which can lead to sodium and water retention and exacerbate the symptoms of heart failure.

In addition, thyroid hormones have been shown to directly affect the expression of genes involved in cardiac remodeling and fibrosis. In particular, excess thyroid hormone has been associated with increased expression of genes involved in collagen synthesis and deposition, which can lead to the development of myocardial fibrosis and impaired cardiac function.

Overall, the underlying mechanisms by which thyroid hormones influence the prognosis of heart failure are complex and multifaceted. Further research is needed to fully elucidate these mechanisms and to identify potential therapeutic targets for improving the outcomes of patients with heart failure.

## Limitations of the study

This study has several limitations. It was a retrospective study and subject to biases inherent in such studies. Future directions for research Future research should explore the mechanisms by which FT3 affects the prognosis of patients with heart failure. Additionally, prospective studies with larger sample sizes and multiple centers are needed to validate our findings and determine the optimal cut-off value for FT3 in identifying high-risk patients. Finally, interventional studies are needed to investigate the effect of interventions to optimize FT3 levels on the prognosis of patients with heart failure.

## Clinical recommendations

Based on our findings, it is recommended that clinicians assess FT3 levels in Hypertrophic cardiomyopathy patients with HFpEF, in addition to other established prognostic factors such as age, atrial fibrillation, and NT-proBNP. Patients with low FT3 levels should be closely monitored for adverse outcomes and may benefit from targeted interventions to improve their thyroid function. Additionally, our study highlights the importance of identifying and addressing potentially modifiable factors, such as thyroid dysfunction, in the management of Hypertrophic cardiomyopathy patients with HFpEF.

## Conclusion

In conclusion, our study found that FT3 is an independent prognostic factor in Hypertrophic cardiomyopathy patients with HFpEF, and the non-linear predictive value of FT3 for prognosis is presented in an “L” shape, with a critical value of FT3=2.885. These findings are important for clinicians to identify high-risk patients and adjust treatment strategies accordingly. These findings have important implications for identifying high-risk patients who may benefit from early intervention and monitoring. Clinicians should consider measuring FT3 levels in patients with heart failure and monitoring them closely for adverse outcomes. Future research should explore the underlying mechanisms linking FT3 levels and adverse cardiovascular outcomes and evaluate the effectiveness of interventions aimed at improving thyroid hormone levels in patients with heart failure.

However, this study has several limitations, including the relatively small sample size, the lack of information on potential confounding factors, and the limited generalizability of the results. Future studies with larger sample sizes, more comprehensive data, and more diverse populations are needed to further validate our findings and explore the underlying mechanisms. Overall, our study highlights the importance of considering FT3 as a prognostic factor in Hypertrophic cardiomyopathy patients with HFpEF and provides new insights into the management of this condition.

## Consent for publication

All authors have approved the final manuscript for publication.

## Availability of data and materials

Not applicable.

## Author Contributions

X.B conducted statistical analyses and collaborated on drafting the manuscript. X.B, J.G, K.Z, W.X.L, W.Y.W, C.S and Y.D.T reviewed and edited the manuscript and contributed to the discussion. W.Y.W, C.S and Y.D.T. obtained funding, conceived the research, and drafted the manuscript. All authors approved the final version of the manuscript. X.B is the guarantor of this work and, as such, had full access to all of the data in the study and takes responsibility for the integrity of the data and the accuracy of the data analysis.

## Acknowledgments

The authors thank all the investigators and subjects who participated in this project.

